# Race/ethnicity in relation to incident primary open-angle glaucoma characterized by autonomously determined visual field loss patterns

**DOI:** 10.1101/2021.10.25.21265498

**Authors:** Jae H. Kang, Mengyu Wang, Lisa Frueh, Bernard Rosner, Janey L. Wiggs, Tobias Elze, Louis R. Pasquale

## Abstract

**Purpose:** We used an autonomous algorithm to classify incident visual field (VF) loss patterns in primary open-angle glaucoma (POAG). Subsequently, we compared racial differences in the risk of these regional VF loss patterns.

**Design/Participants:** Participants (n=209,036) from the Nurses’ Health Study (NHS) (follow-up: 1980-2018); NHS2 (1989-2019); and Health Professionals Follow-up Study (HPFS; 1986-2018), aged ≥40 years and free of glaucoma.

**Methods:** Demographics, medical and lifestyle information was assessed on biennial questionnaires. Incident POAG cases (n=1946) with reproducible Humphrey VF loss were confirmed with medical records. The total deviation information of the earliest reliable VF for each eye with POAG (n=2564) was extracted, and a statistical learning method was used to identified optimal solutions for regional vision loss patterns. Each POAG eye was assigned the VF pattern (“archetype”) based on the highest weighting coefficient.

Multivariable-adjusted hazard ratios (HRs) for POAG of various archetypes and 95% confidence intervals (CIs) were estimated using per-eye Cox proportional hazards models. Covariates included cohort, age, glaucoma family history, socioeconomic status, lifestyle parameters, number of eye exams during follow-up, and medical conditions. False discovery rate (FDR) was used for multiple comparisons.

**Main outcome measures:** POAG based on VF patterns.

**Results:** Mean age was 58 years; 1.3% were Black, 1.2% were Asian, 1.1% were Hispanic-White and 96.4% were non-Hispanic White. We identified 14 archetypes: 1 representing no VF loss, 9 of early loss and 4 of advanced loss patterns. Compared to non-Hispanic Whites, Blacks were at significantly higher risk of POAG with early VF loss archetypes collectively (Blacks: HR=1.96, 95% CI=1.46, 2.63) and at even higher risk for POAG with advanced loss archetypes collectively (Blacks: HR=6.07, 95% CI=3.61, 10.21; p=0.0002 for the two estimates being different); no differences were observed for Asians or Hispanic Whites. For individual VF archetypes, Hispanic-Whites had FDR-significant higher risks of POAG of archetypes showing early paracentral defects and advanced superior loss while Blacks had FDR-significant higher risks of all advanced loss archetypes and 3 early loss patterns, including early paracentral defects.

**Conclusion:** Among health professionals, compared to non-Hispanic-Whites, Blacks and Hispanic-Whites had higher risks of incident POAG with central and advanced VF loss.

---

Primary open-angle glaucoma (POAG) is a complex, multifactorial chronic optic neuropathy that manifests as distinct visual field (VF) loss patterns localizing to the nerve fiber layer.^1^ Previous studies have manually documented patterns of new onset of glaucomatous VF loss among patients with ocular hypertension, and from such studies, it is clear that multiple distinct loss patterns exist,^2, 3^ suggesting that both the patterns of underlying optic nerve damage and the etiology in POAG are heterogenous.^4, 5^ In contrast to evaluating ‘all POAG’ or POAG stratified by intraocular pressure (IOP) levels, studies of POAG incorporating the heterogeneity in VF loss patterns representing different types of optic nerve damage may provide new etiologic insights.

Automated VF data is a spatial array of retinal sensitivities reflecting the functional integrity of the entire visual pathway.^6^ VF mean deviation (MD), pattern standard deviation (PSD) and the glaucoma hemifield test represent useful indices, but they provide no information regarding the regional nature of VF loss.^7, 8^ Archetype analysis is an artificial intelligence (AI) algorithm that analyzes data that clusters on the edges of data space for ascertaining dimensional patterns in a dataset.^9^ When applied to Humphrey VF data from a tertiary care glaucoma clinic, archetype analysis objectively identified weighted patterns of VF loss that were strikingly similar to manually documented VF patterns for patients with new-onset POAG.^2, 10^ The weighting coefficients derived from archetype analysis can help establish an earlier glaucoma diagnosis^11^ and aid in determining and quantifying glaucomatous VF progression.^12^

We applied archetype analysis to new onset POAG in 3 prospective US population-based health professional cohorts to ascertain VF loss patterns. Then, because early disease tends to be asymmetric, we assessed the inter-eye correlation between patterns of VF loss.^13, 14^ Finally, given that African heritage is a strong POAG risk factor,^15-17^ we evaluated whether Blacks, Asians and Hispanic Whites were predisposed to specific VF loss patterns.

## METHODS

### Study population

The Nurses’ Health Study (NHS) began in 1976 when 121,700 female nurses aged 30–55 years were recruited. The NHS2 was initiated in 1989 with 116,429 female nurses aged 25–42 years. The Health Professionals Follow-up Study (HPFS) enrolled, in 1986, 51,529 male health professionals aged 40–75 years. Since the initial recruitment health questionnaires, biennial follow-up surveys have been administered to collect information on lifestyle, diet, and medical status, including information about physician-diagnosed glaucoma. A total of 209,036 participants from the NHS (N=79,895; follow-up period: 1980-2018), NHS2 (N=86,795; follow-up period: 1989-2019) and the HPFS (N=42,346; follow-up period: 1986-2018) were included. We excluded participants with prevalent glaucoma and prevalent cancer (as cancer profoundly changes lifestyle), those without a baseline food frequency questionnaire (FFQ) in NHS and HPFS (as dietary exposures were of main interest in the initial glaucoma studies and thus those without baseline FFQs were not followed) and those who only completed the baseline (1980/1989/1986) questionnaires and were lost to follow-up. Follow-up response rates have been >85%. The institutional review boards of the Brigham and Women’s Hospital, Harvard T.H. Chan School of Public Health and Icahn School of Medicine at Mount Sinai approved the study protocol; participants’ completion of questionnaires were considered implied consent by the IRBs. This study adhered to the tenets of the Declaration of Helsinki.

### Assessment of race/ethnicity and potential risk factors for POAG

We used participants’ self-reported information on biennial questionnaires for covariates potentially related to POAG in prior studies (**eMethods1**): age, race, ethnicity, socioeconomic status, glaucoma family history, body mass index (BMI), mean arterial blood pressure, hypertension, diabetes mellitus, hypercholesterolemia, myocardial infarction, total cholesterol level, physical activity, cigarette smoking, beta-blocker and other anti-hypertensives use, statin and other cholesterol lowering drug use, healthy eating index, dietary intakes of caffeine, alcohol, and nitrate, markers of access to eye care (e.g., self-reports of cataract, cataract extraction, age-related macular degeneration, and number of eye exams), number of other physician visits and among women, age at menopause and postmenopausal hormone use. Validation studies have found a high reliability and accuracy of information from our health professional participants.^18^ If missingness was <5%, values were imputed to the median (for continuous variables); if missingness was greater, missingness indicators were created for covariates.

### Assessment of POAG cases and extraction of VF data

When participants reported new-onset glaucoma on biennial questionnaires, we asked them for permission to obtain confirmatory medical data from their eye care providers. We obtained medical records or a completed glaucoma questionnaire with items including maximal IOP, filtration apparatus status, optic nerve structural information, ophthalmic surgery, and all VF data. Then, a glaucoma specialist (LRP) reviewed the medical records to confirm a diagnosis of POAG using standardized criteria.

For POAG confirmation, we required: (a) gonioscopy indicating the trabecular meshwork was visible in both eyes (70% of cases) or slit lamp biomicroscopy demonstrating normal anterior chamber depth plus pharmacological dilation (30% of cases); (b) slit lamp biomicroscopy demonstrating no signs in either eye of pigment dispersion syndrome, uveitis, exfoliation syndrome, trauma, or rubeosis; and (c) reproducible VF defects consistent with glaucoma on ≥2 reliable tests. The type of perimetry was restricted to 24-2 or 30-2 Humphrey VFs performed with full thresholding or the Swedish Interactive Thresholding Algorithm strategy.

A total of 1957 participants **(eTable1**) were diagnosed with incident POAG (NHS: 1251 cases, NHS2: 223 cases, HPFS: n=483 cases). In eyes with POAG (n=2581), the total deviation (dB) values from the earliest VF test indicating glaucomatous loss were extracted, and VF loss patterns were determined. For those with bilateral POAG, the worse eye was defined as the one with the lower MD value; for those with unilateral POAG, the worse eye was the eye with POAG, and data from the non-affected eye was not used. The median time between the date of first glaucomatous sign (IOP>21mmHg; cup disc ratio (CDR)>0.6 or asymmetry>0.1; or evidence of glaucomatous VF loss) and the VF test in the worse eye was 1 year, and this did not differ by race (p>0.10).

### Statistical Analyses

#### Determining Archetypal VF Loss Patterns

Archetypal analysis (an unsupervised AI technique) on extracted total deviation (dB) data, was applied to determine VF loss patterns for each POAG affected eye. Archetypal analysis reduces dataset dimensionality by anchoring datapoints to values on the edges of a data cluster, autonomously generating and quantifying VF patterns that are clinically recognizable, valid and useful^11, 12, 19^ (**eMethods2 and eFigure1** for an example).

#### Inter-eye analysis of archetypal VF loss patterns

There were 624 pairs of eyes with bilateral POAG available to conduct inter-eye association analyses. We calculated the inter-eye Spearman correlations for the weighting coefficients of the archetypal VF loss patterns between the worse and better eyes. P-values were corrected for multiple comparisons using false discovery rate (FDR).^20^ We further evaluated the relation between the weighting coefficients of the archetypal VF loss patterns in the worse eye with the weighting coefficients of each archetypal VF loss pattern in the better eye using the stepwise Bayesian information criterion (BIC) method. The statistical importance of each parameter was measured by the magnitude of BIC increase when a parameter was removed from the optimal model. When the BIC increase for a parameter was at least 6 higher than another parameter in the model, the former parameter was considered more strongly associated with the outcome than the latter parameter.^9^

#### Prospective per-eye analysis of race/ethnicity of POAG subtypes defined by archetypes

For the prospective analysis, to maximize power and as early POAG can be asymmetrical, we used the eye as the unit of analysis adjusting for inter-eye correlations, with “eye-years” accrued over time as has been previously described.^21, 22^ For each eye with POAG, the archetype with the highest weighting coefficient was used for assigning POAG subtypes based on regional VF loss (**eFigure1)**. For eyes where the highest weighting coefficient was for the normal VF pattern (Figure 1; archetype 1), we assigned the archetype with the second highest weighting coefficient. The diagnosis date was the earliest date of any glaucomatous sign (IOP, CDR, or VF loss) in either of the affected eye(s); we stopped follow-up at this date to minimize incorporating post-diagnosis changes in covariates. For each eye, eye-years of follow-up were accrued from the return of the baseline questionnaire until glaucoma diagnosis, cancer, loss to follow-up, death, or study completion, whichever came first.

**Figure 1.**
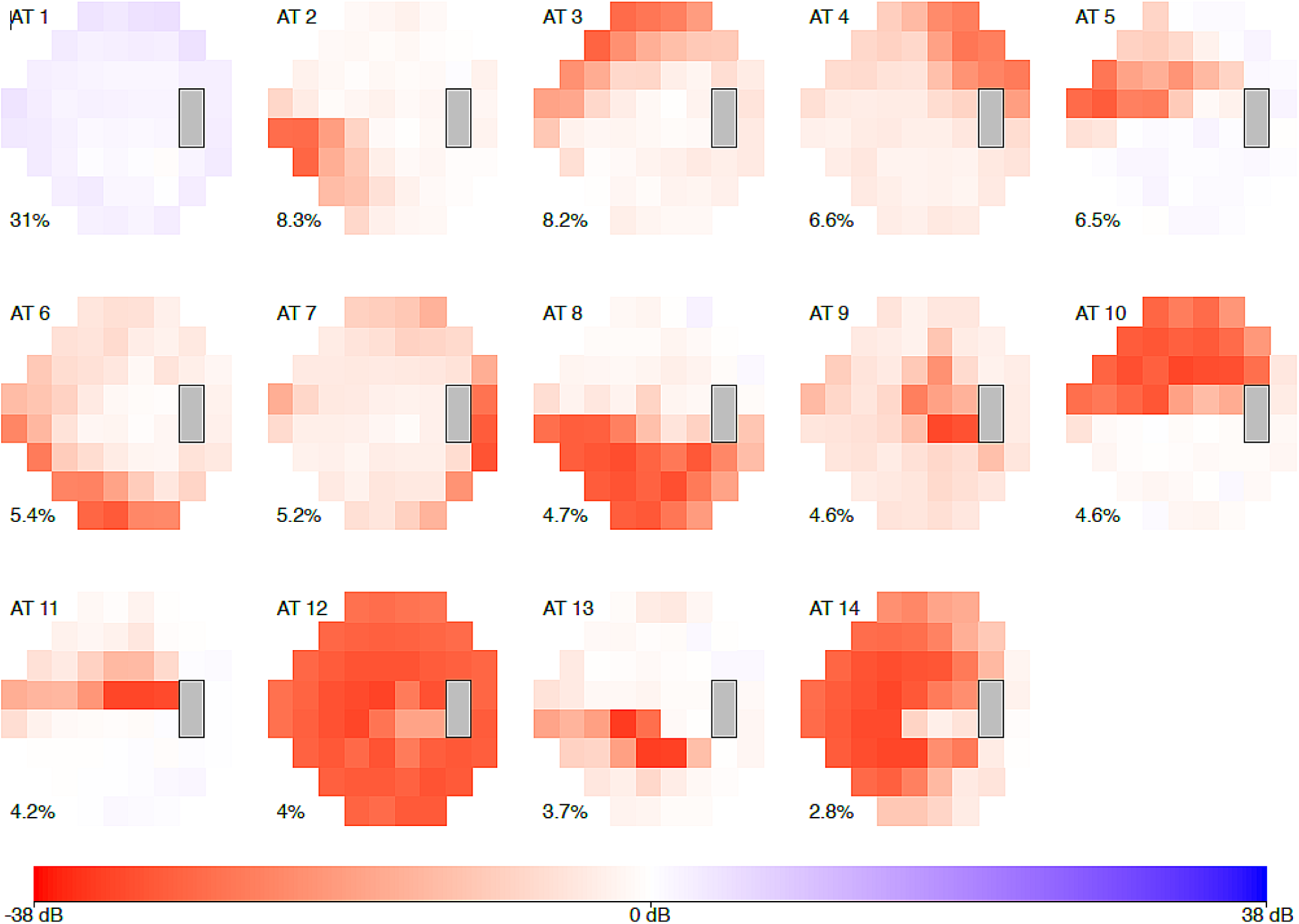
The 14 archetypal visual field loss patterns (ATs) derived from visual fields of the 1957 incident primary open-angle glaucoma cases (2581 affected eyes). The integer at the top left of each archetype denotes the archetype number. The percentage at the bottom left of each archetype indicates the respective average decomposition weight for this pattern.

We combined the data from our three cohorts, then evaluated per-eye Cox proportional hazards models^21, 22^ using age as the time metameter with time-varying covariates that stratified on age in months,^23^ 2-year risk period and cohort, adjusting for the correlation of VF loss in the 2 eyes, to estimate multivariable-adjusted hazard ratios (HRs) and 95% confidence intervals (CIs). Analyses were performed with SAS 9.4 (SAS Institute, Cary NC). For associations with individual POAG subtypes defined by VF archetypes, p<0.05 based on FDR^20^ was considered statistically significant to address multiple comparisons. We used the contrast test method^24^ to evaluate whether the association with at least one archetype was different from the others.

## RESULTS

### Determining archetypal VF loss patterns and inter-eye analysis of archetypal VF loss patterns

Archetype analyses identified 14 archetypal VF loss patterns (AT) in 2581 eyes with incident POAG (**Figure 1**). AT 1 (normal VF pattern) was the most prevalent AT, followed by patterns resembling superior (AT 2) and inferior nasal steps (AT 3). Most patterns resembled pathology affecting the retinal nerve fiber layer except for AT 4 and 9, which were possibly non-glaucomatous VF loss patterns.

In inter-eye analyses **(Figure 2)**, the highest Spearman correlation coefficients between the weighting coefficients in the worse (horizontal axis) and better (vertical axis) eyes were found between the same archetypal VF loss patterns (r range: 0.13–0.63, p<0.003). Comparable results were observed in stepwise regression analyses that evaluated the relation between the archetypal VF loss patterns in the worse eye with each of the 14 archetypal VF loss patterns in the better eye (**eFigure2**) or when instead of worse-better eye comparisons, we evaluated correlations **(eFigure3)** or regression analyses **(eFigure4)** between the right and left eyes.

**Figure 2.**
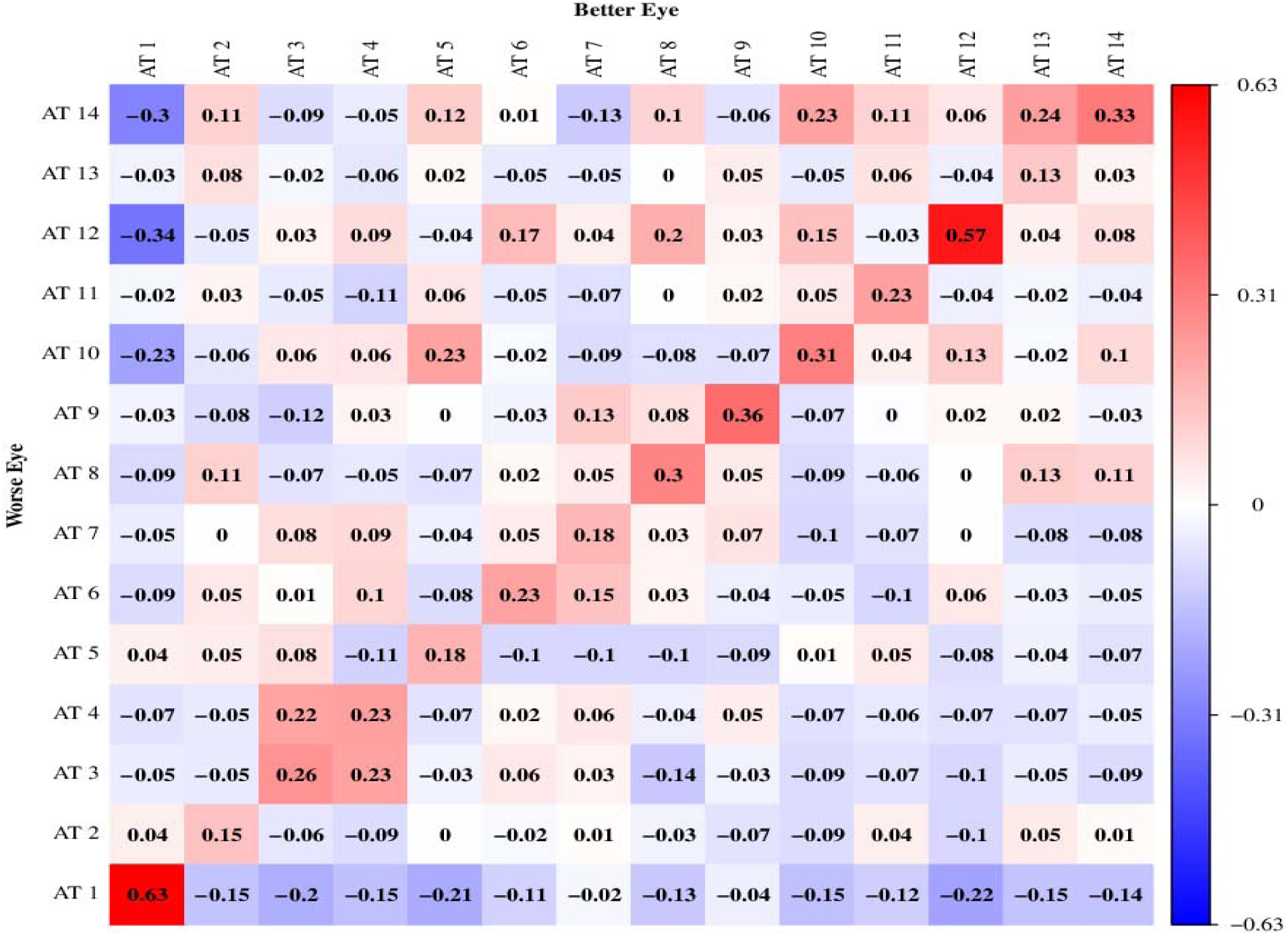
Spearman correlation coefficients between the weight coefficients of the 14 archetypal VF loss patterns in the better (vertical axis) and worse (horizontal axis) eyes among 624 incident primary open-angle glaucoma cases who were affected in both eyes. Blue and red denote positive and negative correlations, respectively.

### Per eye prospective analysis of race/ethnicity of POAG subtypes defined by archetypes

For the per-eye analyses of POAG subtypes defined by VF archetypes, we censored 10 cases as they developed cancer during follow-up and 1 case whose highest weighting coefficient was for the normal VF pattern and did not have a second highest coefficient. This left 2564 eyes with VF loss from 1946 incident POAG cases (1250 NHS cases, 216 NHS2 cases, 480 HPFS cases) for analyses.

Compared to Non-Hispanic Whites, Blacks were younger and more frequently reported a glaucoma family history. Blacks had more diabetes, hypertension, higher BMI, lower socioeconomic status than Non-Hispanic Whites, and among women, they were less likely to take postmenopausal hormones (**Table 1**). Compared to Non-Hispanic Whites, Asians were younger, had more diabetes, more hypertension, and lower BMI. Asians smoked and drank alcohol less than Non-Hispanic Whites, and among women, were less likely to take postmenopausal hormones. Hispanic Whites were younger than Non-Hispanic Whites, had more frequent family history of glaucoma, more diabetes, but smoked less. Overall, Blacks and Asians had the fewest eye and physician exams (**Table 1**). Among cases (**eTable2**), Black and Hispanic White POAG cases were the youngest at diagnosis and were the most likely to have both eyes affected while Asian POAG cases had the lowest IOP and highest CDR.

**Table 1.** Age and age-adjusted characteristics of “eye-years (ey)” of follow-up by race / ethnicity in the Nurses’ Health Study (1980-2018), Nurses’ Health Study II (1989-2019) and Health Professionals Follow-up Study (1986-2018)^a^

|  | Non-Hispanic<br>-White<sup>b</sup><br>(ey=7,638,258) | Black<sup>b</sup><br>(ey=100,022) | Asian<sup>b</sup><br>(ey=97,943) | Hispanic-White<sup>b</sup><br>(ey=86,617) |
| --- | --- | --- | --- | --- |
| % of total person-time | 96.4 | 1.3 | 1.2 | 1.1 |
| Age, years (SD) | 58.1 (11.2) | 56.8 (10.6) | 56.8 (10.9) | 55.8 (10.5) |
| Female, % | 83.8 | 91.3 | 78.6 | 89.7 |
| Family history of glaucoma, % | 18.6 | 28.9 | 18.3 | 24.9 |
| Self-reported diabetes, % | 6.3 | 12.9 | 9.7 | 9.8 |
| Self-reported hypertension, % | 34.7 | 52.4 | 39.0 | 36.0 |
| Self-reported cataract diagnosis, % | 14.6 | 14.6 | 14.3 | 15.3 |
| Self-reported cataract extraction, % | 7.8 | 6.1 | 7.6 | 7.6 |
| Self-reported age-related macular degeneration, % | 2.7 | 1.7 | 2.1 | 2.2 |
| Mean body mass index, kg/m<sup>2</sup> (SD) | 25.4 (4.7) | 27.6 (5.3) | 23.6 (3.5) | 25.9 (4.7) |
| Mean physical activity, MET-hours/week (SD) | 21.2 (22.6) | 18.0 (22.7) | 21.3 (24.1) | 21.7 (24.0) |
| Mean pack-years of smoking (SD) | 9.5 (19.4) | 6.7 (16.0) | 4.6 (18.3) | 5.8 (17.1) |
| Mean caffeine intake, mg / day (SD) | 261.5 (199.7) | 172.2 (157.0) | 203.9 (175.5) | 227.1 (178.1) |
| Mean alcohol intake, g / day (SD) | 6.1 (9.3) | 3.1 (6.2) | 2.8 (6.8) | 4.6 (7.5) |
| Mean AHEI score (without alcohol) (SD) | 47.3 (9.7) | 49.1 (10.1) | 51.2 (9.7) | 50.1 (9.6) |
| Mean age at menopause, years (SD)<sup>c</sup> | 49.1 (4.8) | 48.8 (4.9) | 49.5 (4.2) | 48.9 (4.7) |
| Current postmenopausal hormone use, <sup>c</sup> % | 20.8 | 14.4 | 17.8 | 20.6 |
| Mean # eye exams reported during follow-up (SD) | 5.3 (4.1) | 4.6 (3.9) | 4.9 (4.1) | 5.2 (4.2) |
| Mean # physician visits reported during follow-up (SD) | 7.9 (3.8) | 7.2 (3.8) | 7.5 (3.8) | 8.3 (3.6) |
| Socioeconomic status score based on census tract<sup>d</sup> (SD) | 0.2 (4.7) | -5.2 (6.6) | 0.6 (5.3) | -0.6 (5.6) |

Abbreviations: AHEI = Alternate Healthy Eating Index score (without alcohol; range 0-100); ey = eye-years of follow-up; MET-hours = metabolic equivalent-hours; SD = standard deviation.
<sup>a</sup> Values are means (SD) or percentages and are standardized to the age distribution of the study population.
<sup>b</sup> Due to the small categories and for simplicity, those self-reporting any African ancestry were first categorized as Blacks, then among those remaining, those self-reporting any Asian ancestry were categorized as Asians, then among those remaining, those self-reporting Hispanic ethnicity were categorized as Hispanic-White; all others were categorized as non-Hispanic-White.
<sup>c</sup> Among women only.
<sup>d</sup> This score is based on the sum of the z-scores of census tract indicators based on participants’ zip codes (median household income, home value, percentage with college degree, percentage of families with interest or dividends, percentage occupied housing, percentage living in poverty, percentage White).

Of the 13 ATs showing loss, four (AT 8, 10, 12, and 14) represented “advanced VF loss”, while the other 9 were considered “early VF loss” (**Figure 1**). Compared to Non-Hispanic Whites, Blacks were significantly more likely to develop POAG with early VF loss, with the various nested models showing similar associations (**Table 2**: Model 3: Blacks: HR=1.96, 95% CI=1.46, 2.63); while for POAG with advanced VF loss, Blacks were at even higher risk (Blacks: HR=6.07, 95% CI=3.61, 10.21). Notably, the difference in the associations for POAG with advanced VF loss versus POAG with early VF loss in Blacks versus Non-Hispanic Whites, was statistically significant (p for difference in estimates=0.0002); the elevated risks were not different for the two subtypes of POAG for Asians (p=0.81) or Hispanic-Whites (p=0.46). Indeed, in multivariable-adjusted linear regression analyses of MD in POAG eyes, compared to Non-Hispanic-Whites, Blacks had a significantly worse MD (difference in MD = −2.18; 95% CI=−3.21, −1.15); this was not observed for Asians or Hispanic-Whites (p≥0.23).

**Table 2.**
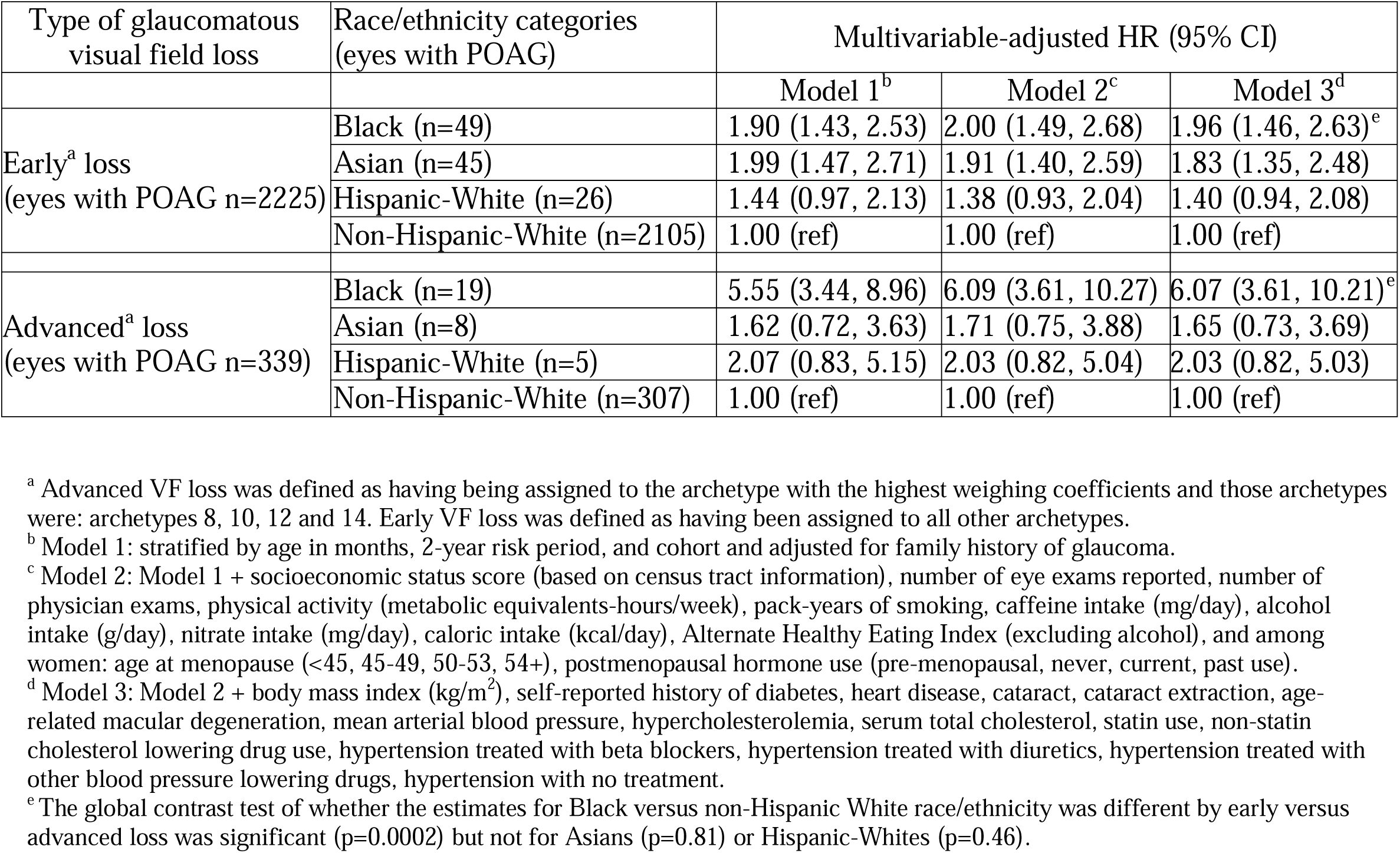
Relative risks of glaucoma with early VF loss versus advanced VF loss archetypes based on the highest weighting coefficients of the affected eye(s)^a^ by race, compared to not developing any glaucoma

When the 13 ATs were evaluated individually by race/ethnicity (**Table 3**), we observed that globally, there were no significant differences in associations for Asians (p=0.87) or Hispanic-Whites (p=0.19) compared to Non-Hispanic Whites; however, we observed globally significant differences (p=0.01) across archetypes for Blacks compared to Non-Hispanic Whites. Specifically, in Model 3, Blacks had FDR-significantly higher risks of developing POAG for 3 of 9 early VF loss archetypes: AT 3 (HR=2.83, 95% CI=1.63, 4.92), AT 5 (HR=2.44, 95% CI=1.16, 5.15), and AT 11 (HR=3.65, 95% CI=1.46, 9.11) and all 4 advanced VF loss archetypes: AT 8 (HR=7.49, 95% CI=3.32, 16.90), AT 10 (HR=3.48, 95% CI=1.16, 10.50), AT 12 (HR=15.06, 95% CI=5.02, 45.18), and AT 14 (HR=6.41, 95% CI=1.56, 26.40). Hispanic Whites had FDR-significantly higher risk of the advanced VF loss archetype, AT10 (HR=4.80, 95% CI=1.71, 13.47), and AT 11 (HR=4.53, 95% CI=1.78, 11.54), consistent with paracentral VF loss.

**Table 3.**
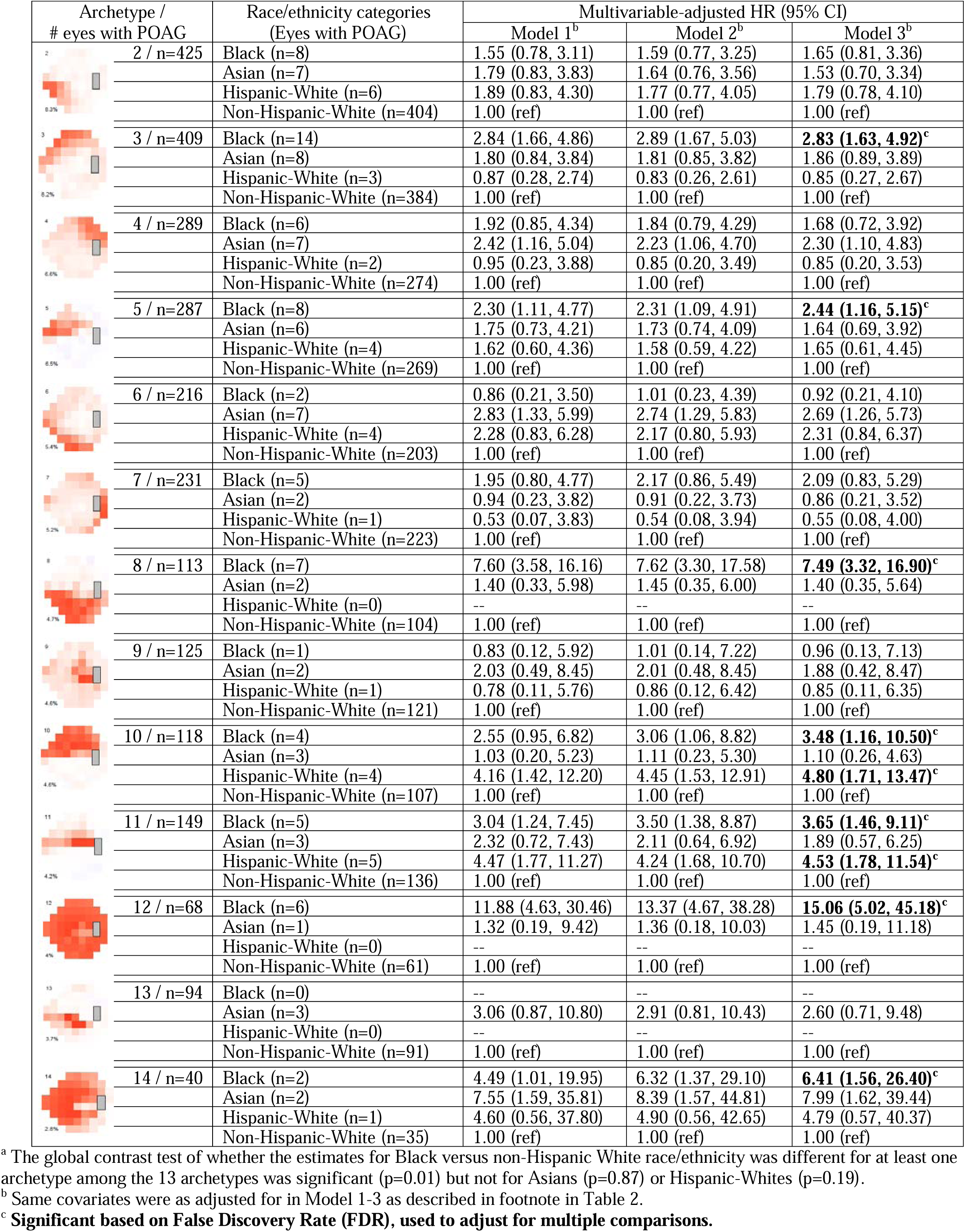
Relative risks for race and archetypes based on the highest weighting coefficients of the eye with POAG^a^

| Archetype /<br># eyes with POAG | Race/ethnicity categories<br>(Eyes with POAG) | Multivariable-adjusted HR (95% CI) |  |  |
| --- | --- | --- | --- | --- |
|  |  | Model 1<sup>b</sup> | Model 2<sup>b</sup> | Model 3<sup>b</sup> |
| 2 / n=425 | Black (n=8) | 1.55 (0.78, 3.11) | 1.59 (0.77, 3.25) | 1.65 (0.81, 3.36) |
|  | Asian (n=7) | 1.79 (0.83, 3.83) | 1.64 (0.76, 3.56) | 1.53 (0.70, 3.34) |
|  | Hispanic-White (n=6) | 1.89 (0.83, 4.30) | 1.77 (0.77, 4.05) | 1.79 (0.78, 4.10) |
|  | Non-Hispanic-White (n=404) | 1.00 (ref) | 1.00 (ref) | 1.00 (ref) |
| 3 / n=409 | Black (n=14) | 2.84 (1.66, 4.86) | 2.89 (1.67, 5.03) | **2.83 (1.63, 4.92)<sup>c</sup>** |
|  | Asian (n=8) | 1.80 (0.84, 3.84) | 1.81 (0.85, 3.82) | 1.86 (0.89, 3.89) |
|  | Hispanic-White (n=3) | 0.87 (0.28, 2.74) | 0.83 (0.26, 2.61) | 0.85 (0.27, 2.67) |
|  | Non-Hispanic-White (n=384) | 1.00 (ref) | 1.00 (ref) | 1.00 (ref) |
| 4 / n=289 | Black (n=6) | 1.92 (0.85, 4.34) | 1.84 (0.79, 4.29) | 1.68 (0.72, 3.92) |
|  | Asian (n=7) | 2.42 (1.16, 5.04) | 2.23 (1.06, 4.70) | 2.30 (1.10, 4.83) |
|  | Hispanic-White (n=2) | 0.95 (0.23, 3.88) | 0.85 (0.20, 3.49) | 0.85 (0.20, 3.53) |
|  | Non-Hispanic-White (n=274) | 1.00 (ref) | 1.00 (ref) | 1.00 (ref) |
| 5 / n=287 | Black (n=8) | 2.30 (1.11, 4.77) | 2.31 (1.09, 4.91) | **2.44 (1.16, 5.15)<sup>c</sup>** |
|  | Asian (n=6) | 1.75 (0.73, 4.21) | 1.73 (0.74, 4.09) | 1.64 (0.69, 3.92) |
|  | Hispanic-White (n=4) | 1.62 (0.60, 4.36) | 1.58 (0.59, 4.22) | 1.65 (0.61, 4.45) |
|  | Non-Hispanic-White (n=269) | 1.00 (ref) | 1.00 (ref) | 1.00 (ref) |
| 6 / n=216 | Black (n=2) | 0.86 (0.21, 3.50) | 1.01 (0.23, 4.39) | 0.92 (0.21, 4.10) |
|  | Asian (n=7) | 2.83 (1.33, 5.99) | 2.74 (1.29, 5.83) | 2.69 (1.26, 5.73) |
|  | Hispanic-White (n=4) | 2.28 (0.83, 6.28) | 2.17 (0.80, 5.93) | 2.31 (0.84, 6.37) |
|  | Non-Hispanic-White (n=203) | 1.00 (ref) | 1.00 (ref) | 1.00 (ref) |
| 7 / n=231 | Black (n=5) | 1.95 (0.80, 4.77) | 2.17 (0.86, 5.49) | 2.09 (0.83, 5.29) |
|  | Asian (n=2) | 0.94 (0.23, 3.82) | 0.91 (0.22, 3.73) | 0.86 (0.21, 3.52) |
|  | Hispanic-White (n=1) | 0.53 (0.07, 3.83) | 0.54 (0.08, 3.94) | 0.55 (0.08, 4.00) |
|  | Non-Hispanic-White (n=223) | 1.00 (ref) | 1.00 (ref) | 1.00 (ref) |
| 8 / n=113 | Black (n=7) | 7.60 (3.58, 16.16) | 7.62 (3.30, 17.58) | **7.49 (3.32, 16.90)<sup>c</sup>** |
|  | Asian (n=2) | 1.40 (0.33, 5.98) | 1.45 (0.35, 6.00) | 1.40 (0.35, 5.64) |
|  | Hispanic-White (n=0) | -- | -- | -- |
|  | Non-Hispanic-White (n=104) | 1.00 (ref) | 1.00 (ref) | 1.00 (ref) |
| 9 / n=125 | Black (n=1) | 0.83 (0.12, 5.92) | 1.01 (0.14, 7.22) | 0.96 (0.13, 7.13) |
|  | Asian (n=2) | 2.03 (0.49, 8.45) | 2.01 (0.48, 8.45) | 1.88 (0.42, 8.47) |
|  | Hispanic-White (n=1) | 0.78 (0.11, 5.76) | 0.86 (0.12, 6.42) | 0.85 (0.11, 6.35) |
|  | Non-Hispanic-White (n=121) | 1.00 (ref) | 1.00 (ref) | 1.00 (ref) |
| 10 / n=118 | Black (n=4) | 2.55 (0.95, 6.82) | 3.06 (1.06, 8.82) | **3.48 (1.16, 10.50)<sup>c</sup>** |
|  | Asian (n=3) | 1.03 (0.20, 5.23) | 1.11 (0.23, 5.30) | 1.10 (0.26, 4.63) |
|  | Hispanic-White (n=4) | 4.16 (1.42, 12.20) | 4.45 (1.53, 12.91) | **4.80 (1.71, 13.47)<sup>c</sup>** |
|  | Non-Hispanic-White (n=107) | 1.00 (ref) | 1.00 (ref) | 1.00 (ref) |
| 11 / n=149 | Black (n=5) | 3.04 (1.24, 7.45) | 3.50 (1.38, 8.87) | **3.65 (1.46, 9.11)<sup>c</sup>** |
|  | Asian (n=3) | 2.32 (0.72, 7.43) | 2.11 (0.64, 6.92) | 1.89 (0.57, 6.25) |
|  | Hispanic-White (n=5) | 4.47 (1.77, 11.27) | 4.24 (1.68, 10.70) | **4.53 (1.78, 11.54)<sup>c</sup>** |
|  | Non-Hispanic-White (n=136) | 1.00 (ref) | 1.00 (ref) | 1.00 (ref) |
| 12 / n=68 | Black (n=6) | 11.88 (4.63, 30.46) | 13.37 (4.67, 38.28) | **15.06 (5.02, 45.18)<sup>c</sup>** |
|  | Asian (n=1) | 1.32 (0.19, 9.42) | 1.36 (0.18, 10.03) | 1.45 (0.19, 11.18) |
|  | Hispanic-White (n=0) | -- | -- | -- |
|  | Non-Hispanic-White (n=61) | 1.00 (ref) | 1.00 (ref) | 1.00 (ref) |
| 13 / n=94 | Black (n=0) | -- | -- | -- |
|  | Asian (n=3) | 3.06 (0.87, 10.80) | 2.91 (0.81, 10.43) | 2.60 (0.71, 9.48) |
|  | Hispanic-White (n=0) | -- | -- | -- |
|  | Non-Hispanic-White (n=91) | 1.00 (ref) | 1.00 (ref) | 1.00 (ref) |
| 14 / n=40 | Black (n=2) | 4.49 (1.01, 19.95) | 6.32 (1.37, 29.10) | **6.41 (1.56, 26.40)<sup>c</sup>** |
|  | Asian (n=2) | 7.55 (1.59, 35.81) | 8.39 (1.57, 44.81) | 7.99 (1.62, 39.44) |
|  | Hispanic-White (n=1) | 4.60 (0.56, 37.80) | 4.90 (0.56, 42.65) | 4.79 (0.57, 40.37) |
|  | Non-Hispanic-White (n=35) | 1.00 (ref) | 1.00 (ref) | 1.00 (ref) |

<sup>a</sup> The global contrast test of whether the estimates for Black versus non-Hispanic White race/ethnicity was different for at least one archetype among the 13 archetypes was significant (p=0.01) but not for Asians (p=0.87) or Hispanic-Whites (p=0.19).
<sup>b</sup> Same covariates were as adjusted for in Model 1-3 as described in footnote in Table 2.
<sup>c</sup> Significant based on False Discovery Rate (FDR), used to adjust for multiple comparisons.

## DISCUSSION

Using an unsupervised AI algorithm, we identified 14 ATs in incident POAG from three population-based cohorts. In case-only analyses, in general, the best predictor of the weighting coefficients of each archetype in the better eye were those of the same archetype in the worse eye. Also, while recognizing that race is an inexact proxy for multiple attributes including cultural, societal, environmental, biological and other factors,^25^ we observed that even after adjusting for many factors, compared with non-Hispanic Whites, Black and Hispanic White participants were at significantly increased risk of POAG with advanced and central VF loss. This is notable given that our participants were health professionals with high levels of education and similar access to healthcare.

The ATs observed were like those generated by Elze et al.^10^ in a tertiary glaucoma clinic. Both studies found that the normal VF pattern was most common, that superior and inferior nasal steps were common early defects and that both solutions autonomously recognized a dense superior paracentral VF loss pattern. Like Teng et al.,^26^ we found a strong inter-eye association between in the patterns of VF loss, indicating the within-person consistency and possibly implicating systemic susceptibilities caused by genetics and environmental exposures shared between eyes of the same patient.

Other population-based POAG studies have observed a higher prevalence, earlier POAG onset and more severe VF loss at diagnosis in Blacks and among Hispanics.^15, 27-31^ These findings may be due to less access to or utilization of eye care, higher prevalence of risk factors, genetic differences, chronic stress or a combination of factors. One proposed explanation for racial/ethnic health disparities is that minorities experience higher allostatic load (i.e., physiological burden of stress measured using biomarkers pertinent to cardiovascular, metabolic, inflammatory, and neuroendocrine systems^32^) and health deterioration earlier in life than non-Hispanic-Whites due to the cumulative impact of marginalization and discrimination, a concept known as ‘weathering’.^33^ While our multivariable-adjusted model adjusted for several of these downstream biomarkers (i.e., age, diabetes, blood pressure), stress and inflammatory biomarkers related to discrimination that we did not account for may have contributed to higher incidence of glaucoma and greater glaucoma disease severity at diagnosis.^34^

Eye care utilization differs among racial / ethnic groups, with Blacks being least likely to have regular eye exams in the U.S.^35^ However, given that our cohort consists of health professionals, and that we allowed in analyses only those who reported eye exams in the past 2 years and that racial / ethnic differences were observed even after adjustment for number of eye exams during follow-up, it is unlikely that eye care access differences drove the racial/ethnic differences observed.

Genetic factors may also have played a role, as Black and Hispanic White participants had comparatively more frequent glaucoma family history. Genetically determined African ancestry has been independently associated with greater glaucoma risk,^36^ and among Blacks, family history was associated with an earlier age at diagnosis.^37^ Also, in Latinos, having more African ancestry informativity genetic markers was associated with higher IOP.^38^ Thus, in Black and Hispanic White population, earlier and more frequent eye exams may have public health importance.

A strength of our study was the use of a novel archetype analysis to generate quantitative measures of regional patterns of VF loss. This was a large prospective study with 1946 incident cases (2564 eyes with POAG) and 209,036 participants followed for 30+ years, with high follow-up rates. With a wealth of information, particularly repeated health and behavior information and markers of socioeconomic status^30^ and the homogeneity of the study population in education and health care access, we were able to minimize the possibility of confounding biases.

Our study had several limitations. Repeated in-person eye exams were not possible, and thus, we relied on questionnaire and medical record information for disease confirmation, a method that had low sensitivity. However, methodologically, hazard *ratios* can still be valid if the case definition is highly specific (e.g., reproducible VF loss) and the ascertainment method was unrelated to exposure (we required reports of eye exams at each follow-up cycle).^39^ Furthermore, on all participants, we did not have regularly updated information on IOP information and central corneal thickness (CCT). Yet, CCT is not considered a strong POAG risk factor^40^ in the general population, and in the Baltimore Eye Study (and among our cases; **eTable2**), IOP was similar in prevalent POAG cases among Blacks and Whites.^15^ Finally, because our study participants were health professionals, our results may not be generalizable to general populations, where racial/ethnic disparities in POAG may be larger.

In summary, in this prospective study of incident POAG among health professionals, archetype analyses were able to identify and quantify major specific regional patterns of VF loss, and when compared to non-Hispanic-Whites, Blacks and Hispanic-Whites, had higher risks of incident POAG with central and advanced loss.

## Data Availability

Information including the procedures to obtain and access data from the Nurses Health Studies and Health Professionals Follow-up Study is described at https://www.nurseshealthstudy.org/researchers (contact) and https://sites.sph.harvard.edu/hpfs/for-collaborators/

## SUPPLEMENTAL MATERIAL

### Supplemental Methods text 1

#### eMethods1. Assessment of race/ethnicity and potential risk factors for POAG

Race and ethnicity was assessed in 1992 and 2004 in NHS; 1989 and 2005 in NHS2 and 1986 and 2014 in HPFS. Due to the small categories and for simplicity, those self-reporting any African ancestry were first categorized as Blacks, then among those remaining, those self-reporting any Asian ancestry were categorized as Asians, then among those remaining, those self-reporting Hispanic ethnicity were categorized as Hispanic-White; all others were categorized as non-Hispanic-White. Age was calculated as years from birthdate until the return of each questionnaire. Cigarette smoking details were assessed biennially, and pack-years of smoking was updated. Participants’ reported time spent per week on 8-10 selected activities was multiplied by each activity’s energy expenditure requirements (metabolic equivalents [METs]) and summed to yield MET-hours-per-week.^1,2^ Weight was assessed biennially, and along with height (assessed in 1976 in NHS, 1989 in NHS2 and 1986 in HPFS), was used to calculate updated cumulatively averaged BMI (kg/m^2^). With cumulative averaging, every two years, the average of all available information was used (e.g., in 1986, the 1986 BMI values were used; in 1988, the average of 1986 and 1988 values was used; in 1990, the average of 1986, 1988 and 1990 values were used, etc.). This approach was used because glaucoma is a chronic disease, and cumulative averages represent participants’ long-term exposure; also, with this approach, no participants had missing data. Diet was assessed in 1980 in NHS, 1991 in NHS2 and 1986 in HPFS and every 2-4 years thereafter using semi-quantitative food frequency questionnaires (SFFQs). In SFFQs, participants reported the frequency of average consumption of a portion size of specific foods or beverages (e.g., beer, wine, spirits, coffee, tea, cola)^3^ during the previous year. Nutrient values were calculated by multiplying the consumption frequency of each food / beverage portion by the nutrient content, summing these products across all items, and then adjusting for total energy intake,^4^ and cumulatively averaged. Self-reported history of diabetes, systolic and diastolic blood pressure, total serum cholesterol, hypercholesterolemia heart disease, hypertension and various types of cholesterol and blood-pressure-lowering medications were repeatedly asked in questionnaires. Mean arterial blood pressure was derived by using the updated cumulatively averaged values for systolic blood pressure and diastolic blood pressure in the following equation: [(1/3*systolic blood pressure) + (2/3*diastolic blood pressure)].

## Supplemental Methods text 2

### eMethods2

#### Archetypal analysis

The optimal number of archetypal VF loss patterns were determined with 10-fold cross-validation by minimizing the overall VF reconstruction errors.^1^ In particular, the data were randomly divided into 10 subsets, where each of the 10 subsets was used as the testing set once, with the remaining nine subsets used as the training set. The VF reconstruction errors on the test set were calculated for the number (**k**) of archetypes from 2 to 20 (the potential range of number of archetypes). The VF reconstruction errors were calculated as the differences between the original VFs and the reconstructed VFs, which were the sum of the archetypes multiplied by their linear decomposition coefficients. The optimal pattern number **k**_**o**_ was determined by the following criteria: the minimum **k** with reconstruction errors that are not statistically different (i.e., Bayes factor < 3.0)^2^ from the **k** that produced the lowest average reconstruction error. Once the optimal number of patterns **k**_**o**_ was determined, the archetypal VF loss patterns were determined over all data using that number. After the archetypal VF loss patterns were determined, each VF was decomposed as a linear combination of the archetypal VF loss patterns with decomposition coefficient adding up to 100%.

## Supplemental Table 1

**eTable 1.**
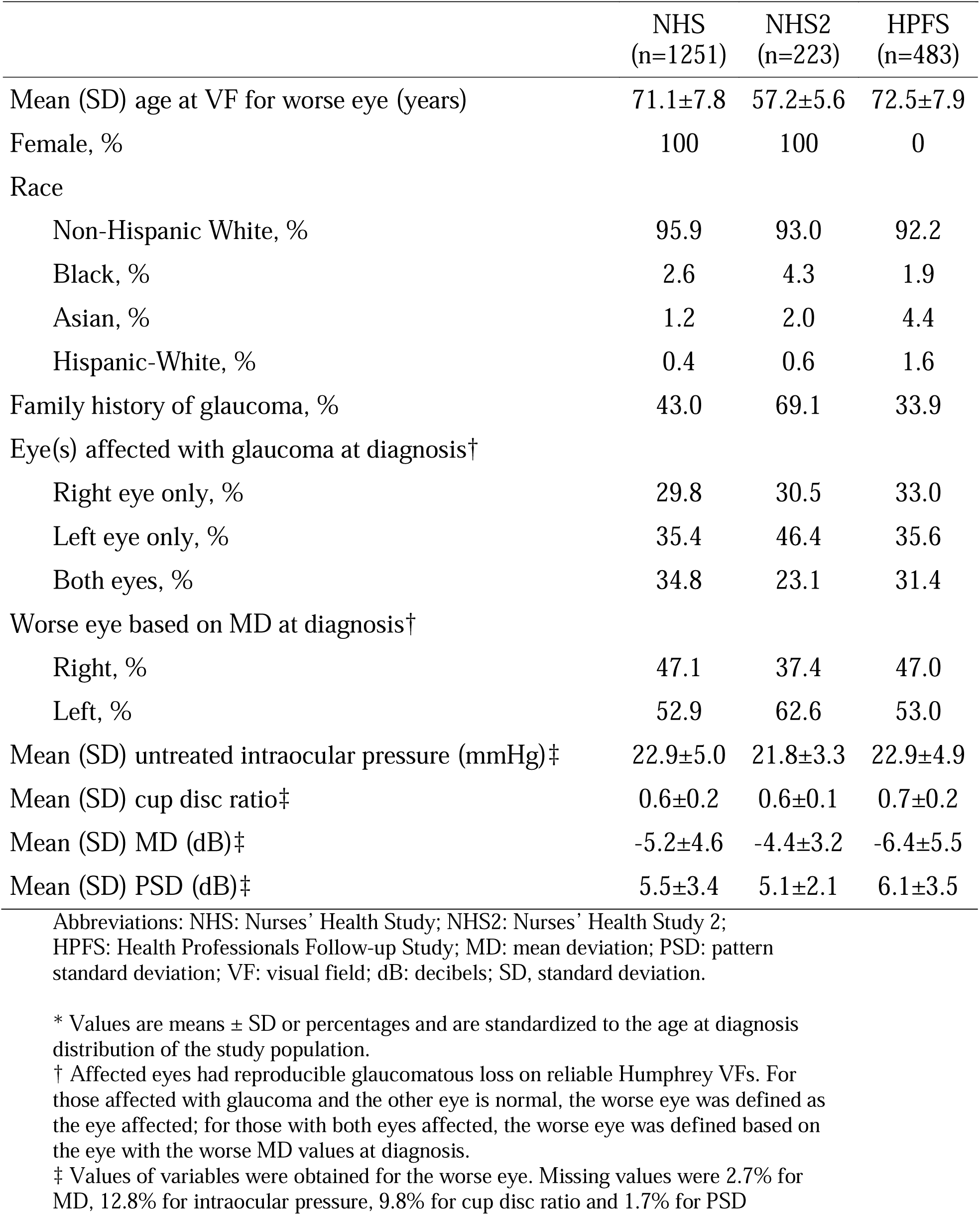
Characteristics of Incident Primary Open-Angle Cases for Archetype analyses in

## Supplemental Table 2

**eTable 2.** Age and age-adjusted characteristics of incident POAG cases (n=1946) and affected eyes (n=2564) as of diagnosis by race / ethnicity in the Nurses’ Health Study (1980-2018), Nurses’ Health Study II (1989-2019) and Health Professionals Follow-up Study (1986-2018)^a^

|  | Non-Hispanic-White<sup>b</sup> | Black<sup>b</sup> | Asian<sup>b</sup> | Hispanic-White<sup>b</sup> |
| --- | --- | --- | --- | --- |
| *Person-level characteristics* | N=1836 | N=50 | N=39 | N=21 |
| % | 94.3 | 2.6 | 2.0 | 1.1 |
| Age at diagnosis, years (SD) | 67.0 (9.4) | 64.5 (8.4) | 66.1 (8.1) | 59.6 (9.5) |
| Female, % | 75.6 | 77.5 | 55.3 | 77.6 |
| Family history of glaucoma, % | 33.3 | 36.6 | 40.2 | 33.0 |
| History of diabetes, % | 7.9 | 16.6 | 16.4 | 23.1 |
| History of hypertension, % | 44.6 | 62.8 | 71.4 | 44.1 |
| Mean body mass index, kg/m<sup>2</sup> (SD) | 25.1 (4.1) | 27.0 (6.0) | 24.3 (3.3) | 27.3 (6.9) |
| Mean physical activity, MET-hours/week (SD) | 21.4 (21.4) | 18.8 (21.5) | 34.9 (51.8) | 13.5 (14.8) |
| Mean pack-years of smoking (SD) | 10.5 (18.1) | 6.1 (11.3) | 9.1 (18.4) | 4.6 (8.9) |
| Mean caffeine intake, mg / day (SD) | 268.7 (192.3) | 169.9 (149.3) | 241.6 (187.6) | 204.4 (180.6) |
| Mean alcohol intake, g / day (SD) | 7.3 (10.4) | 2.9 (4.3) | 3.6 (7.7) | 6.8 (10.2) |
| Mean AHEI score (without alcohol) (SD) | 48.4 (9.6) | 48.2 (10.8) | 50.9 (7.0) | 50.5 (8.6) |
| Mean age at menopause at <45 years of age, %<sup>d</sup> | 9.3 | 5.6 | 3.5 | 18.9 |
| Current postmenopausal hormone use, %<sup>d</sup> | 30.7 | 21.6 | 31.9 | 22.5 |
| Self-reported cataract diagnosis, % | 26.9 | 31.5 | 36.7 | 14.2 |
| Self-reported cataract extraction, % | 12.6 | 7.3 | 11.9 | 5.4 |
| Self-reported age-related macular degeneration, % | 5.7 | 7.3 | 0.0 | 0.0 |
| Socioeconomic status score based on census tract (SD)<sup>e</sup> | 0.1 (4.8) | -5.0 (8.5) | 0.5 (4.7) | 0.6 (3.7) |
| Eye(s) affected with glaucoma at diagnosis |  |  |  |  |
| Right eye only, % | 30.6 | 25.8 | 24.2 | 21.9 |
| Left eye only, % | 36.3 | 33.7 | 36.2 | 19.2 |
| Both eyes, % | 33.0 | 40.5 | 39.6 | 59.0 |
| Worse eye based on VF MD at diagnosis |  |  |  |  |
| Right, % | 46.1 | 49.9 | 43.6 | 79.4 |
| Left, % | 53.9 | 50.1 | 56.4 | 20.6 |
| *Eye-level characteristics* | N=2412 | N=68 | N=53 | N=31 |
| Mean intraocular pressure, mmHg (SD)<sup>c</sup> | 22.8 (5.0) | 23.2 (5.2) | 21.3 (5.1) | 24.5 (5.5) |
| Mean cup disc ratio (SD)<sup>c</sup> | 0.6 (0.2) | 0.6 (0.2) | 0.7 (0.2) | 0.6 (0.2) |
| Mean MD, dB (SD)<sup>c</sup> | -5.0 (4.6) | -7.6 (6.6) | -6.7 (5.3) | -5.2 (4.0) |
| Mean PSD, dB (SD)<sup>c</sup> | 5.3 (3.3) | 6.4 (3.9) | 6.1 (3.0) | 5.5 (3.6) |

Abbreviations: MD = mean deviation; PSD = pattern standard deviation; SD = standard deviation
<sup>a</sup> Values are means (SD) or percentages and are standardized to the age distribution of the study population.
<sup>b</sup> See footnote<sup>b</sup> in Table 1.
<sup>c</sup> Based on the eye with the worse MD.
<sup>d</sup> Among women only.
<sup>e</sup> See footnote<sup>d</sup> in Table 1.

## Supplemental Figure 1

**eFigure 1.**
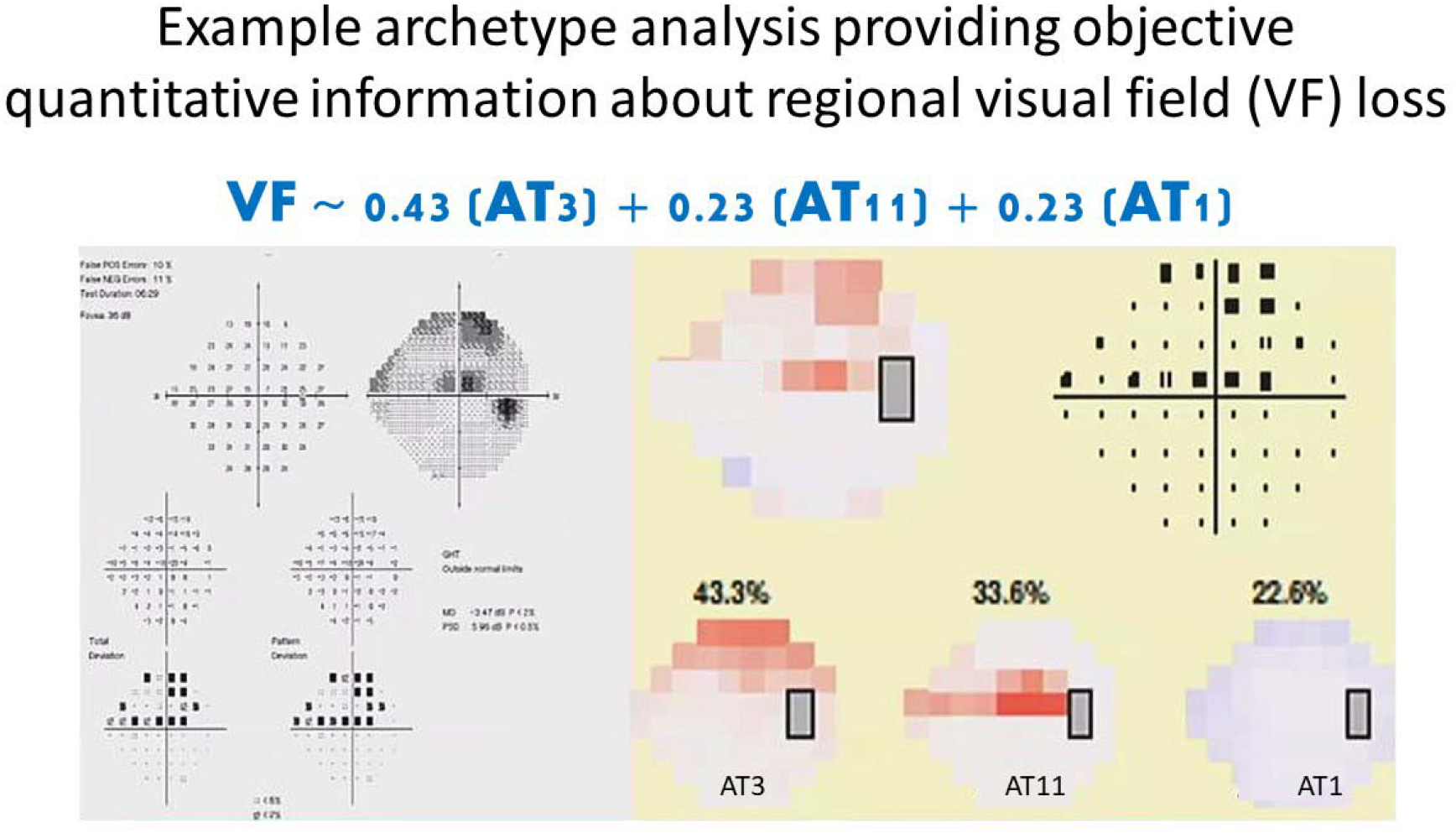
Example showing the input and output of an archetype analysis for a visual field (VF) test (left hand side). The lower right-hand side shows the decomposition of this VF into 3 archetypes (archetypes 3, 11 and 1), with the highest weight coefficient being for archetype 3 (0.43). This POAG affected eye was assigned as having POAG with a VF loss pattern most consistent with archetype 3 in the analyses of the relation between race/ethnicity and POAG subtypes.

## Supplemental Figure 2

**eFigure 2.**
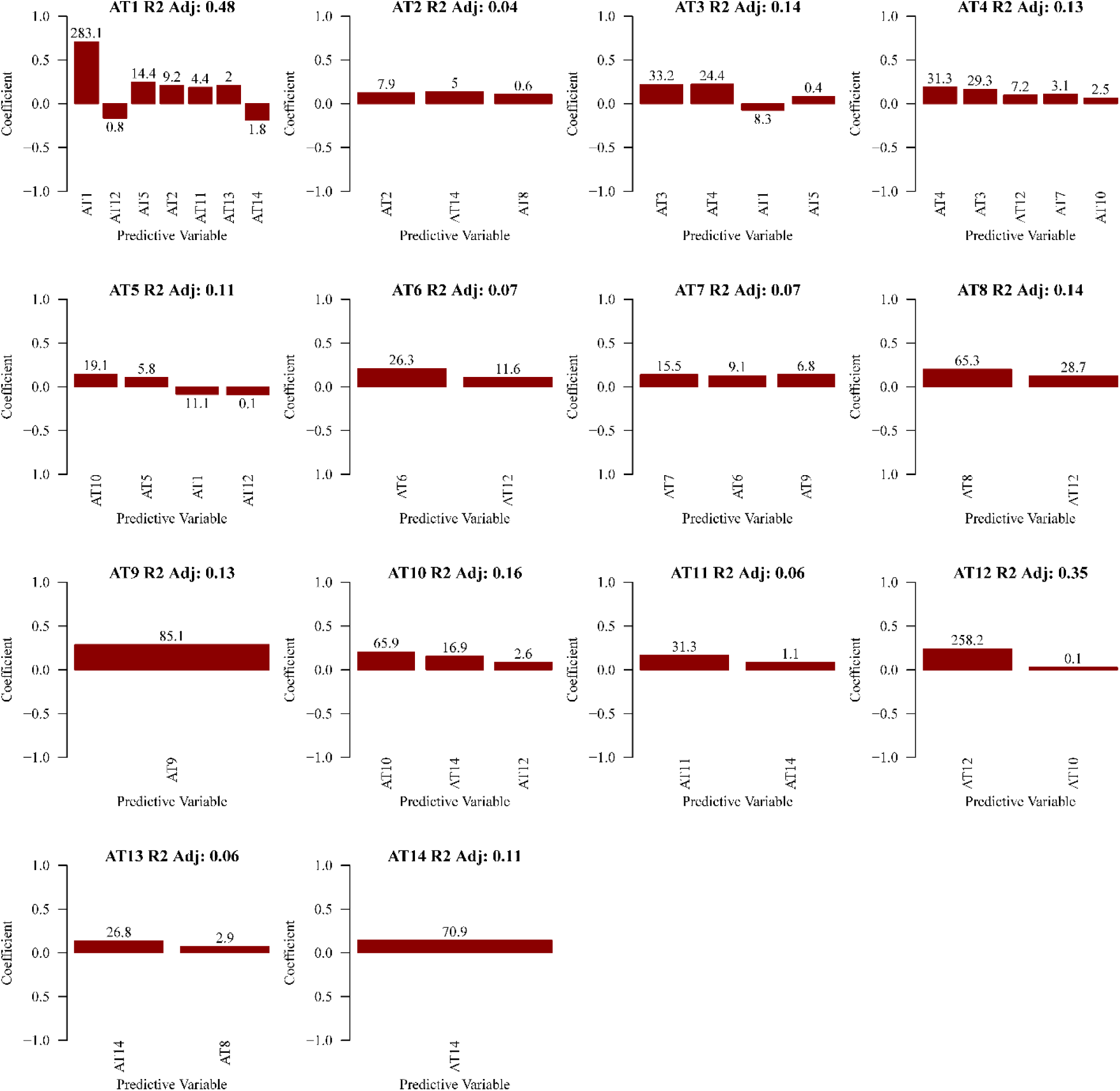
The optimal models to associate the archetypal visual field (VF) loss patterns in the worse eye with each of the 14 archetypal VF loss patterns in the better eye (n=624 pairs). Stepwise regression was applied to remove redundant features; the y-axis represents the betas of the association between parameters from regression models. The R-squared (R^2^) adjusted for the number of parameters was reported to measure the optimal model’s performance. The statistical importance of each parameter was measured by the magnitude of Bayesian information criterion (BIC) increase when a parameter was removed from the optimal model. When the BIC increase for a parameter is at least 6 higher than another parameter in the model, the former parameter is considered strongly more associated with the outcome than the latter parameter.^9^ The adjusted R^2^ values ranged from 0.04 (archetype 2 in the better eye, inferonasal loss) to 0.48 (archetype 1 in the better eye, normal VFs). The five archetypes in the better eye with highest adjusted R^2^ values were: archetype 1 (the normal VF, R^2^ = 0.48), archetype 12 (near total loss, R^2^=0.35), archetype 10 (superior altitudinal loss, R^2^=0.16), AT 8 (inferior altitudinal loss, R^2^=0.14) and AT 3 (superior nasal loss, R^2^=0.14). In general, the same archetype in the worse eye was most predictive of the archetype in the better eye in terms of statistical importance measured by BIC increase except for ATs 5 (superior nasal-paracentral loss) and 13 (inferior paracentral loss) in the better eye. ATs 5 and 13 in the better eye were most positively associated with archetypes 10 (superior altitudinal loss) and 14 (diffuse loss with temporal island) in the worse eye, respectively.

## Supplemental Figure 3

**eFigure 3.**
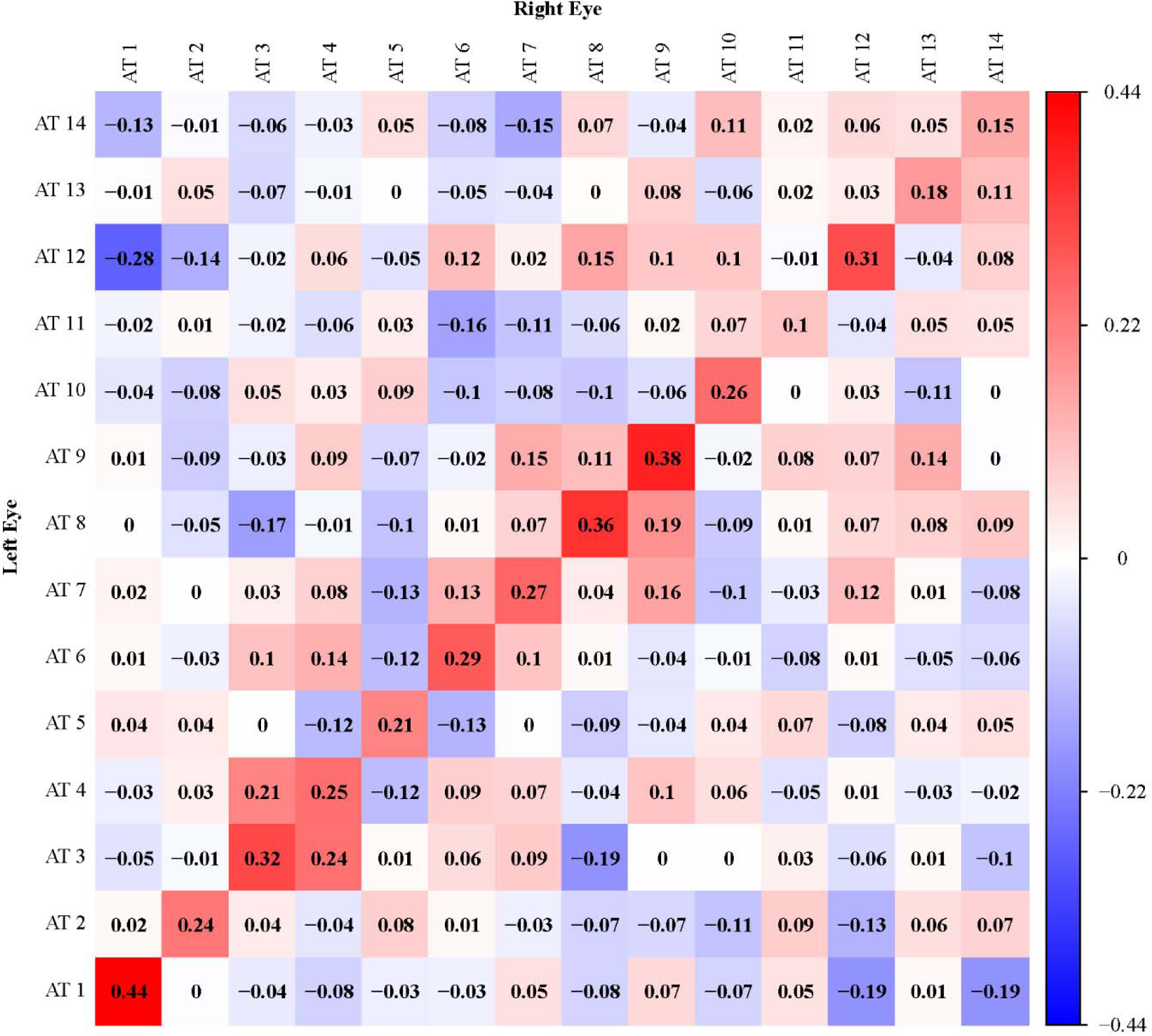
Spearman correlation coefficients between the weight coefficients of the 14 archetypal VF loss patterns in the left eye (vertical axis) and right (horizontal axis) eyes among 624 incident primary open-angle glaucoma cases who were affected in both eyes. Blue and red denote positive and negative correlations, respectively.

## Supplemental Figure 4

**eFigure 4.**
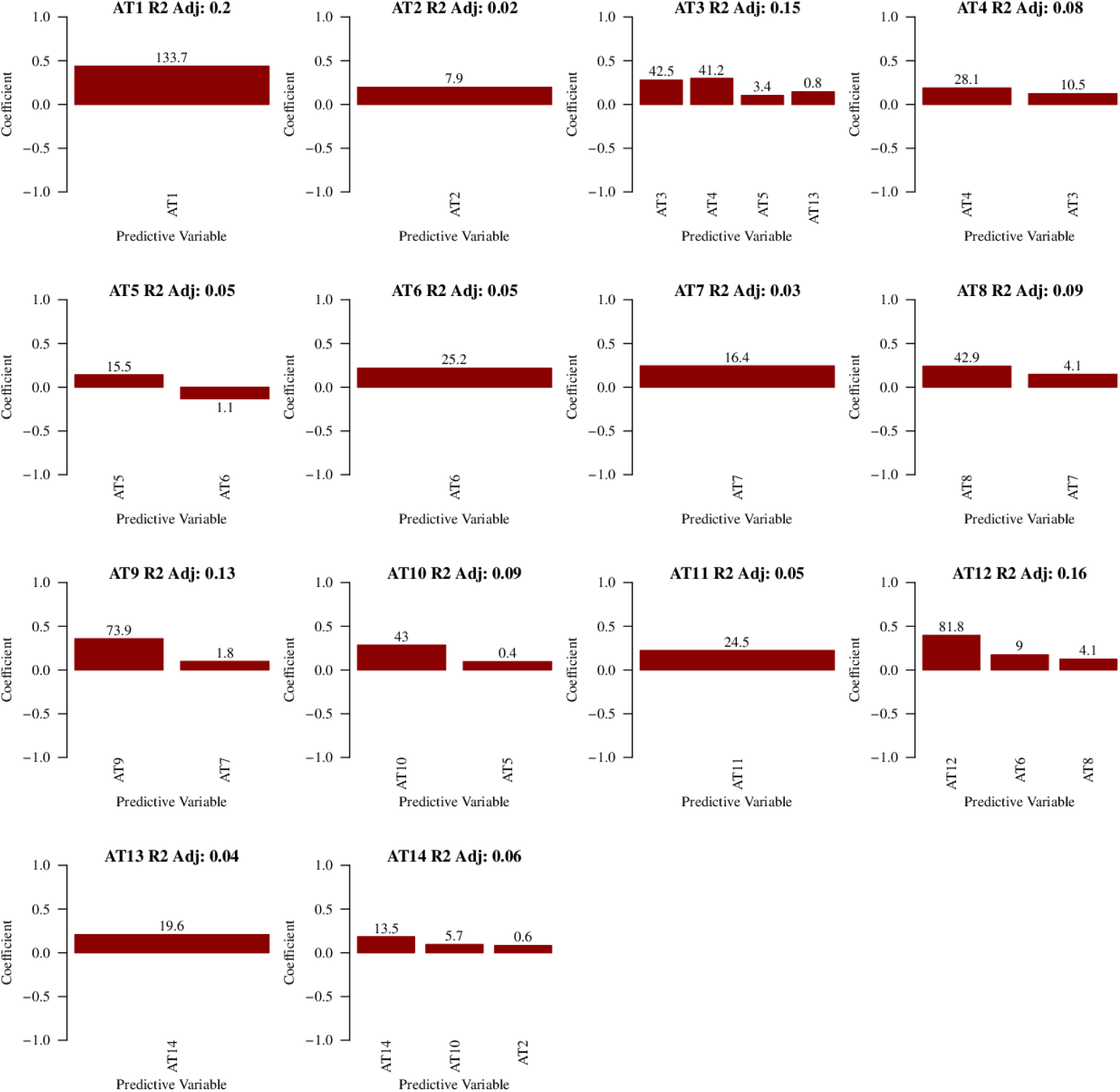
The optimal models to associate the archetypal visual field (VF) loss patterns in the left eye with each of the 14 archetypal VF loss patterns in the right eye (n=624 pairs). Stepwise regression was applied to remove redundant features; the y-axis represents the betas of the association between parameters from regression models. The R-squared adjusted (R^2^) for the number of parameters was reported to measure the optimal model’s performance. The statistical importance of each parameter was measured by the magnitude of Bayesian information criterion (BIC) increase when a parameter was removed from the optimal model. When the BIC increase for a parameter is at least 6 higher than another parameter in the model, the former parameter is considered strongly more associated with the outcome than the latter parameter.^9^ The adjusted R ^2^ values ranged from 0.02 (archetype 2 in the left eye, inferonasal loss) to 0.2 (archetype 1 in the better eye, normal VFs). The six archetypes in the left eye with highest adjusted R^2^ values were: archetype 1 (the normal VF, R^2^= 0.20), archetype 12 (near total loss, R^2^=0.16), AT 3 (superior nasal loss, R^2^=0.15), archetype 9 (central loss, R^2^=0.13), AT 8 (inferior altitudinal loss, R^2^=0.09) and AT 10 (superior altitudinal loss, R^2^=0.09). In general, the same archetype in the right eye was most predictive of the archetype in the left eye in terms of statistical importance measured by BIC increase except for AT 13 (inferior paracentral loss) in the left eye. AT 13 in the left eye were most positively associated with archetype 14 (diffuse loss with temporal island) in the right eye.

